# Time for a standardized neurological assessment in acute setting: the modified Neurological Impairment scale

**DOI:** 10.1101/2025.02.16.25322368

**Authors:** Alessandro Padovani, Irene Mattioli, Tiziana Comunale, Nicola Zoppi, Cinzia Zatti, Enis Guso, Marcello Catania, Andrea Morotti, Chiara Agosti, Stefano Gipponi, Lynne Turner-Stokes, Andrea Pilotto, the School of Neurology Working Group

## Abstract

**Background:** Given the increasing diversity among neurological patients, standardized protocols are essential for evaluating the severity and complexity of the variety of conditions. Aim of the present work was to standardize the assessment of the severity and complexity of neurological impairment in an acute setting by using a modified version of the Neurological Impairment Scale (mNIS).

**Methods:** consecutively hospitalized neurological inpatients underwent a multidimensional standardized assessment of multimorbidity, frailty, functional dependency, and neurological impairment using mNIS and other validated scales. Inter-rater reliability of the mNIS total and sub-scores was evaluated. Construct validity was assessed separately in patients with Cerebrovascular disease performing correlations between corresponding sub-scores of mNIS, original NIS, and National Institutes of Health Stroke Scale (NIHSS). mNIS Complexity Index (mNIS-CI) for neurological severity was used to classify patients into subtle, mild, moderate, and severe impairment.

**Results:** one thousand eighty-one neurological patients admitted to a neurological ward from the emergency setting were enrolled. The inter-rater reliability was remarkable for mNIS total and sub-scores (ICC 0.90, 95% CI 0.82–0.95). The mNIS showed strong construct validity for total and sub-scores compared to other clinical scales (r 0.47-0.97, p<0.001) and 52.7% of patients scored at least one in one of the four newly listed items. The stratification of patients according to mNIS-CI exhibited high construct validity distinguishing the extent of impairment and involved domains.

**Conclusions:** The mNIS is valuable for measuring neurologic severity and complexity in acute inpatients and holds significant potential for application in different settings.

**What is already known on this topic:** Standardization of neurological assessment and development of functional rating scales of global impairment are pivotal to characterize and follow up patients both in the clinical practice and in the research setting.

**What this study adds:** mNIS is a valuable tool to measure neurologic severity and complexity in acute inpatients: the four added items are useful for capturing a broader spectrum of signs and symptoms; the severity and complexity scores provide different information about neurological status and domain imapired at individual level.

**How this study might affect research, practice, or policy:** mNIS can be instrumental in grading the severity of neurological conditions, tracking clinical progress, and gauging response to treatments in the acute setting.

## INTRODUCTION

In medicine, standardized assessments with broad applicability and consistency can be instrumental in defining the severity of conditions, tracking clinical progress, and gauging response to treatments (1). Within neurology, the vast array of specialized subspecialties has given rise to numerous distinct scales tailored to classify and score the severity of patients with validity within specific diagnostic contexts (2). Neurological assessments are customized to suit diagnostic categories and clinical scenarios, catering to contexts that range from acute cerebrovascular incidents to chronic evaluation of patients with inflammatory, neurodegenerative, or neuromuscular disorders. Given the increasing diversity among neurological patients in acute and chronic care settings, standardized protocols are essential for evaluating the course of severity and complexity of neurological conditions (3).

The importance of having a tool able to assess neurological disabilities across different domains and settings is critical to address relevant comparisons across various practices, programs, and population demographics. Furthermore, standardized instruments are necessary to identify specific patient vulnerabilities and effectively monitor responses to personalized care management strategies and targeted treatments (4,5).

Functional rating scales typically assess the ability of patients to perform tasks and roles in everyday life and must be valid, reliable, and responsive (6). Functional rating scales can be useful tools in clinical settings (e.g., acute inpatient evaluation, chronic outpatient follow-up evaluation) and clinical trials, being nothing less than outcome measures. One of the most commonly used rating scales in neurology is the National Institutes of Health Stroke Scale (NIHSS) for clinical stroke severity (7). Since its development, the NIHSS has become a key tool in emergency departments and has been used as a standardized outcome measure in ischemic stroke trials (8,9). However, the NIHSS is not applicable across a broader range of acute, subacute, and chronic neurological conditions, and focuses mainly on left hemispheric lesions (e.g. language impairment) (10).

The Neurological Impairment Scale (NIS) (11) was first developed in the 1990s as a measure of the severity of neurological impairment for clinical application across a broad range of neurological disorders and other disabling conditions (e.g. acquired brain injury, trauma, etc). It evolved over two decades and, because of its broad applicability, in 2010 version 8 was adopted as the standardized measure of impairment in the national clinical database for specialized inpatient neurorehabilitation in the United Kingdom (UK Rehabilitation Outcomes Collaborative, https://www.ukroc.org/ukroc/clinical-tools).

Despite formal evidence for the validity and reliability of the NIS as a measure of neurological impairment (11), its application has been limited to neuro-rehabilitation settings in and outside the UK, and its originators acknowledged some gaps in its coverage of some aspects of clinical impairment.

The present prospective study aimed to test the usefulness of a modified version of the NIS (mNIS) to standardize the assessment of severity and complexity of neurological signs in an acute neurological setting, and to examine its construct validity and reliability. To achieve this, we comprehensively stratified individual neurological impairment by adding to the 17-item NIS four additional items (*Cranial nerves, Coordination, Involuntary movements*, and *Gait*) and devising a mNIS Complexity Index (mNIS-CI). The objectives of the study were: (a) to examine mNIS construct validity compared to other validated functional rating scales for total and sub-scores, (b) to determine its inter-rater reliability, and (c) to stratify patients based on their severity and complexity via the mNIS-CI and validate it across different neurological conditions.

## METHODS

### Study design and participants

The study involved adult (age ≥ 18) neurological inpatients consecutively admitted from the Emergency Room to the Neurology Unit of the Spedali Civili Hospital in Brescia (Italy) between January 2023 and December 2023. Demographic, clinical, treatment, and diagnostic data were collected from both printed and electronic medical records using standardized anonymized data collection forms. Premorbid conditions were assessed upon admission using the Cumulative Illness Rating Scale (CIRS) (12). Functional status was evaluated at admission and discharge using the modified Rankin scale (mRS) (13) and the dependency in basic and instrumental activities of daily living (BADL-IADL) (14). A standard neurological and multidimensional examination was conducted for each patient at admission, which included: NIHSS, Glasgow Coma Scale (GCS) (15), Confusion Assessment Method (CAM) (16), and Gait and Stand Tinetti scale (17). Global cognitive assessment was conducted on all patients using the Mini-Mental State Examination (MMSE) during their stay at the hospital (18).

Raters were the resident physicians working in the Neurology Unit in 2023. They conducted a multidimensional evaluation for each patient upon admission, informed by the patient’s clinical complaints, suspected diagnoses, or imaging exam results that prompted the admission. Senior residents provided training on functional scale ratings included in the previously described multidimensional evaluation to junior residents. The data were imputed by resident physicians and quality control was performed by three independent raters (IM, MC, and APi).

The research protocol was approved by the Ethics Committee of the Brescia Hospital, Brescia, Italy (Neuromultibio study, NP 5285). Written informed consent was collected from each participant and the study was conducted in accordance with the declaration of Helsinki.

Patients who did not undergo a complete multidimensional evaluation and did not receive a definite diagnosis at discharge, recorded using the International Classification of Diseases 10th Revision (ICD-10) code, were excluded (Supplementary Figure 1). The diagnostic categories considered in the present work are: Cerebrovascular disease (Ischemic stroke, Transient Ischemic Attack, Intracerebral hemorrhage, Subarachnoid hemorrhage, and other rare diseases), Epilepsy, Headache, Vertigo, Central Nervous System (CNS) neoplasia, Encephalitis/CNS inflammatory disorders, Encephalopathies/Delirium, Neurodegenerative disorders (NDD), Neuromuscular disorders, Functional Neurological Disorders (FND) and Medically Unexplained Symptoms (MUS), Neurosurgical disorders.

### Modified Neurological Impairment Scale (mNIS)

All patients were evaluated using the NIS (version 8), which comprises 17 items (each rated 0–2 or 0–3, with a total score range of 0–50). A description of the scale, together with detailed instructions for rating, is available on the UKROC website (http://www.csi.kcl.ac.uk/ukroc).

The mNIS comprises 17 items of the NIS with the addition of 4 items: *Cranial nerves* (rated 0–3), *Coordination* (rated 0–3), *Involuntary movements* (rated 0–3), and *Gait* (rated 0–5), giving a total score range of 0–64. The administration of the scale requires 20 minutes and should be performed by healthcare professionals with at least a basic knowledge of the neurological examination. The mNIS scoring sheet with the ICD-10 single-item related coding is provided as Supplemental Material (English and Italian versions).

For the mNIS, the Complexity Index (mNIS-CI) was calculated as the number of categories with a score of 2 or greater (i.e. moderate to severe impairment in the specific domain assessed), and in our cohort its values ranged from 0 to 7.

### Statistical analysis

Statistical analyses were performed using the IBM SPSS Statistics Package Version 26 for Mac (Chicago, IL, USA). Statistical significance was set at P <0.05.

#### Internal consistency, construct validity, and inter-rater reliability

Internal consistency and reliability were tested using Cronbach’s alpha coefficient, with an alpha value of ≥ 0.7 being deemed satisfactory.

To evaluate the construct validity of the mNIS in an acute neurological setting, a correlation analysis was conducted between the total mNIS/NIS and NIHSS scores of patients in the Cerebrovascular disease diagnostic category. For items not included in the NIHSS, we assessed their correlation with scores in different scales evaluating severity in the same neurological domain. In particular, *Mental status*, an item encompassing global cognitive evaluation (attention, orientation, memory) was contrasted with the MMSE, and the CAM total scores; the *Gait* item was contrasted with the Tinetti Gait and Balance total score.

Inter-rater reliability tests were performed on a subset of randomly selected patients (n=40) independently evaluated by two raters at admission (IM and TC). The two raters were blinded to each other and scored the mNIS and NIS in the same patient. Cronbach’s alpha and intraclass correlation coefficient (ICC) were calculated for each sub-item; agreement between the two raters was rated and a total alpha>=0.7 was considered satisfactory.

#### mNIS severity grading and complexity evaluation performance

The distribution of the total scores of mNIS and other scales of the multidimensional evaluation were analyzed across different neurological severity groups, defined as 25th percentile ranges of the mNIS total score, and diagnostic categories. We propose a severity grading system as follows: subtle (<25th percentile), mild (25-50th percentile), moderate (50-75th percentile), and severe (75-100th percentile).

Differences in the mNIS Severity Score and in the mNIS Complexity Index, CIRS and age were tested across neurological severity groups with an ANOVA model. Chi square test was performed for sex.

The construct validity of mNIS across neurological severity groups was evaluated using an ANCOVA model adjusted for the effects of age and sex, in particular severity assessed by the NIHSS, the number of patients with abnormal GCS, and disability assessed by the mRS were considered.

Finally, the distribution of different grading of mNIS-CI was qualitatively evaluated within the mNIS-derived neurological severity groups and specific diagnostic categories, to show the ability of mNIS-CI to capture at once differences in severity and complexity of patients’ clinical presentations.

### Data availability

All study data, including raw and analyzed data and materials, will be available from the corresponding authors upon specific request.

## RESULTS

### Study population

Out of 1245 patients admitted, 1081 underwent the complete evaluation with NIS and mNIS, and the complete multidimensional assessment and were thus included in the final analyses (Supplementary Figure 1).

The cohort was highly heterogeneous, including both young adults (mean age 47.3 ± 14.2, n=454) and elderly patients (mean age 77.74±7.00n=627) with a wide range of comorbidities and premorbid frailty and disability (Supplementary Table 1).

The most common diagnosis was Cerebrovascular disease (62.4%), followed by FND or MUS (7.7%), Vertigo (5.8%), and Epilepsy (5.4%). 1081 patients with diagnosis of Cerebrovascular disease had NIHSS, NIS, and mNIS evaluation completed at admission and discharge.

### Construct validity

The distribution of mNIS scores in patients with different neurological diagnoses are shown in Table 1.

**Table 1:**
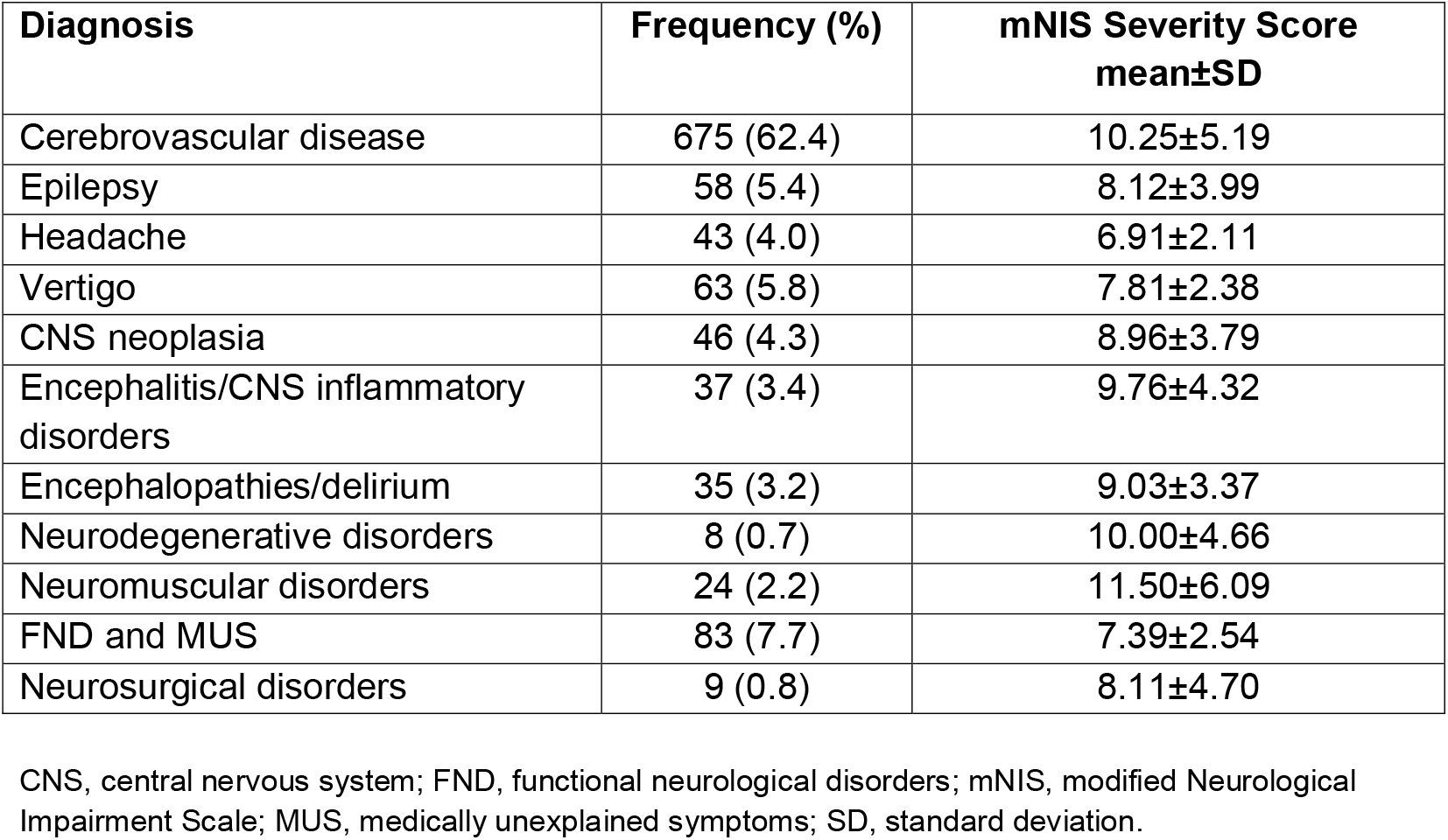
mNIS total score across different neurological conditions.

| Diagnosis | Frequency (%) | mNIS Severity Score<br>mean±SD |
| --- | --- | --- |
| Cerebrovascular disease | 675 (62.4) | 10.25±5.19 |
| Epilepsy | 58 (5.4) | 8.12±3.99 |
| Headache | 43 (4.0) | 6.91±2.11 |
| Vertigo | 63 (5.8) | 7.81±2.38 |
| CNS neoplasia | 46 (4.3) | 8.96±3.79 |
| Encephalitis/CNS inflammatory disorders | 37 (3.4) | 9.76±4.32 |
| Encephalopathies/delirium | 35 (3.2) | 9.03±3.37 |
| Neurodegenerative disorders | 8 (0.7) | 10.00±4.66 |
| Neuromuscular disorders | 24 (2.2) | 11.50±6.09 |
| FND and MUS | 83 (7.7) | 7.39±2.54 |
| Neurosurgical disorders | 9 (0.8) | 8.11±4.70 |

CNS, central nervous system; FND, functional neurological disorders; mNIS, modified Neurological Impairment Scale; MUS, medically unexplained symptoms; SD, standard deviation.

In each diagnostic group, mNIS and NIS showed a high correlation (r ranging from 0.85 to 0.93, p<0.001). Both mNIS and NIS detected neurological impairment in most patients (97.5% and 98.3%, respectively). In the whole group, 52.7% of patients exhibited at least one of four newly listed symptoms, and 33.6% at least two of them. The *Cranial nerve* item was positive in 36.6% of subjects; 19.5% of patients exhibited gait alterations, 3.9% impaired coordination and 3.6% abnormal movements.

The correlation between total mNIS and the NIHSS score was very high in patients with Cerebrovascular disease (r= 0.82, p<0.001, and r= 0.85, p<0.001 respectively). Supplementary Table 2 shows correlation coefficients between mNIS items and the ones measuring the same function/domain from the NIHSS, the Tinetti, the MMSE and the CAM scores, confirming high construct validity for each sub-score included.

### Inter-rater agreement

Supplementary Table 3 reports the level of inter-rater agreement found between mNIS total and sub-scores. Within the two raters, there was high overall agreement for the total NIS score reflected by a Cronbach’s alpha of 0.93 and ICC 0.87 (95% confidence interval 0.77–0.93) and for total mNIS score reflected by a Cronbach’s alpha of 0.95 and ICC 0.90 (95% confidence interval 0.82–0.95). Item-by-item agreement was very good for all subitems of the scale, with lower values for mood, fatigue, and other deficits.

### mNIS score distribution across diagnostic categories and complexity classification

The mNIS total score correlated with all disability measures, including mRS (r=0.31, p<0.001), BADL and IADL (r=0.29, p<0.001 and r=0.27, p<0.001) but not with total comorbidity score assessed by CIRS (p=0.10).

Table 2 shows the distribution of different neurological severity and disability measures according to the mNIS-derived severity classification.

**Table 2.** Demographics and clinical characteristics of patients according to neurological severity.

|  | **Subtle** | **Mild** | **Moderate** | **Severe** | **p** |
| --- | --- | --- | --- | --- | --- |
| Number of patients | 302 | 268 | 283 | 228 |  |
| mNIS score cut-off | 0-5 | 6-8 | 9-12 | $\geq 13$ | |
| mNIS total score, mean±SD | 5.03±1.29 | 7.42±0.50 | 10.19±1.08 | 17.07±3.86 | $p<0.001^{\S}$ |
| mNIS Complexity Index, mean±SD | 0.9±0.3 | 1.4±0.5 | 2.1±0.8 | 3.8±1.3 | $p<0.001^{\S}$ |
| Age, mean±SD | 59.96±19.17 | 60.76±19.57 | 67.47±16.79 | 73.30±13.94 | $p<0.001^{\S}$ |
| Sex, female n% | 53.30 | 42.90 | 48.40 | 55.30 | $p<0.001^*$ |
| CIRS cumulative severity, mean±SD | 1.41±0.35 | 1.49±0.37 | 1.57±0.40 | 1.74±0.49 | $p<0.001^{\P}$ |
| CIRS number of comorbidities 0-1, n% | 51.70 | 43.70 | 34.60 | 21.50 |  |
| 2-3, n% | 31.10 | 35.80 | 38.50 | 35.10 |  |
| 4-6, n% | 16.20 | 17.90 | 23.00 | 32.50 |  |
| > 6, n% | 1.00 | 2.60 | 3.90 | 11.00 |  |
| **Neurological severity measures** |  |  |  |  |  |
| NIHSS total score, n% | 0.86±3.2 | 1.44±1.6 | 3.9±4.1 | 13.1±8.2 | $p<0.001^{\P}$ |
| GCS <15, n% | 0 | 2.6 | 5.8 | 31.2 | $p<0.001^{\P}$ |
| **Disability scales** |  |  |  |  |  |
| mRS premorbid, mean±SD | 0.65±0.89 | 0.86±0.95 | 1.41±1.30 | 2.11±1.62 | $p<0.001^{\P}$ |
| (mRS= 0), n% | 53.30 | 41.00 | 30.70 | 20.20 |  |
| (mRS= 1-2), n% | 41.70 | 52.60 | 47.00 | 39.90 |  |
| (mRS= 3-5), n% | 5.00 | 6.30 | 22.30 | 39.90 |  |

CIRS, Cumulative Illness Rating scale; GCS, Glasgow coma score; mNIS, modified Neurological Impairment Scale; mRS, modified Rankin scale; NIHSS, National Institute of Health Stroke Scale; SD, standard deviation.
§ ANOVA across neurological severity groups
\* Chi square test across neurological severity groups
¶ ANCOVA, covariates: age and sex

The validation of severity classification was supported by the distinct variations observed among the groups about other severity metrics, such as NIHSS (specifically applicable to Cerebrovascular disease diagnosis) and the frequency of patients with abnormal GCS scores (p<0.001). As shown in Figure 1, the total scores of mNIS exhibited variations across various neurological disorders, with Neuromuscular disorders and Headache showing respectively the highest and lowest proportions of more severely affected patients. As shown in Figure 2, the mNIS-CI showed notable variations among neurological severity groups (p<0.001), thereby distinguishing overall severity from the extent of involvement of different neurological domains. Furthermore, substantial variations in mNIS-CI values were observed among different neurological diagnoses, with Encephalitis/CNS inflammatory disorders and Headache presenting respectively the highest and lowest mean mNIS-CI values.

**Figure 1.**
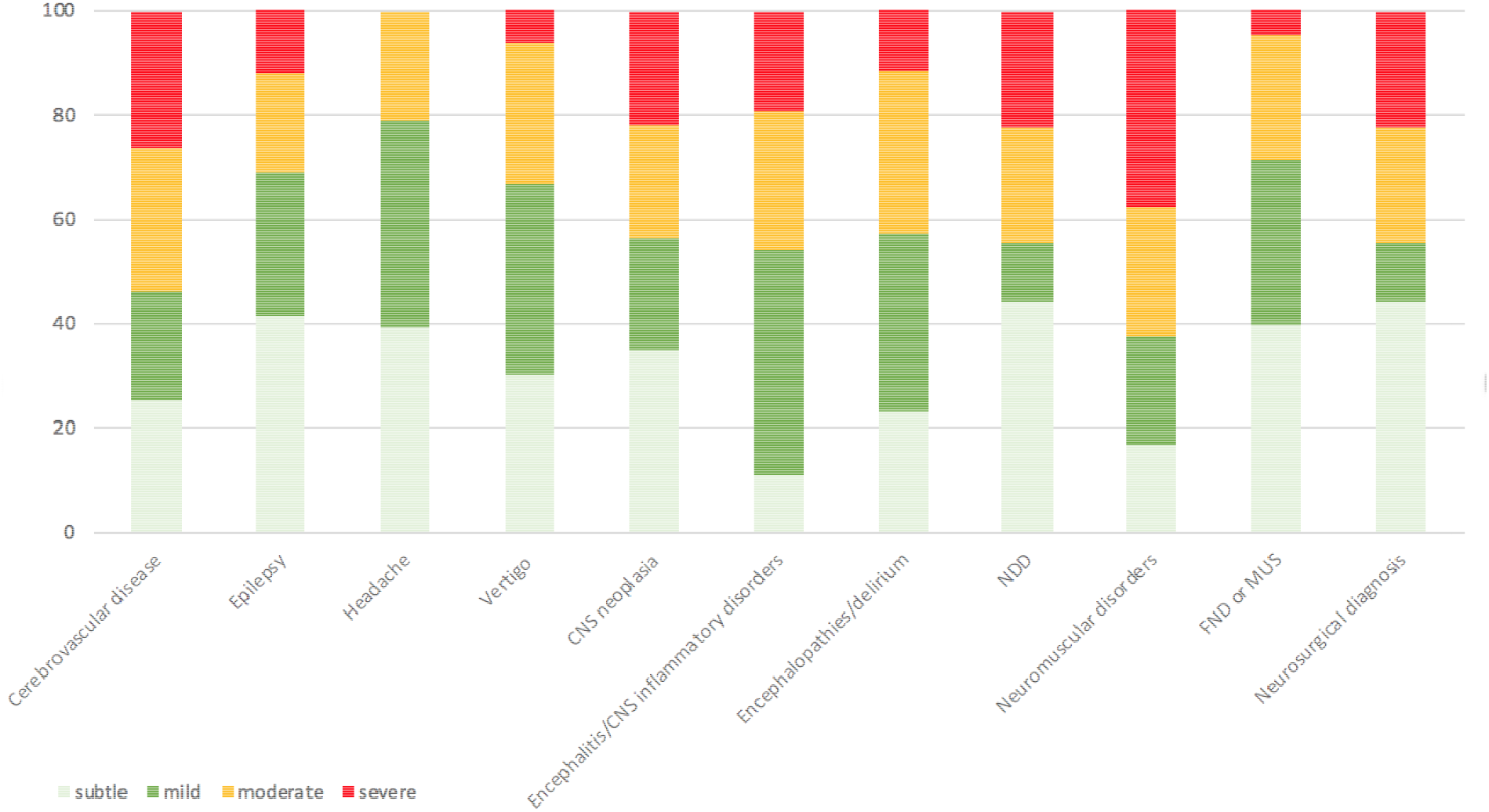
Distribution of neurological severity within neurological disorders: CNS, central nervous system; FND, functional neurological disorders; ICH, intracerebral hemorrhage; MUS, medically unexplained symptoms; NDD, neurodegenerative disorders.

**Figure 2.**
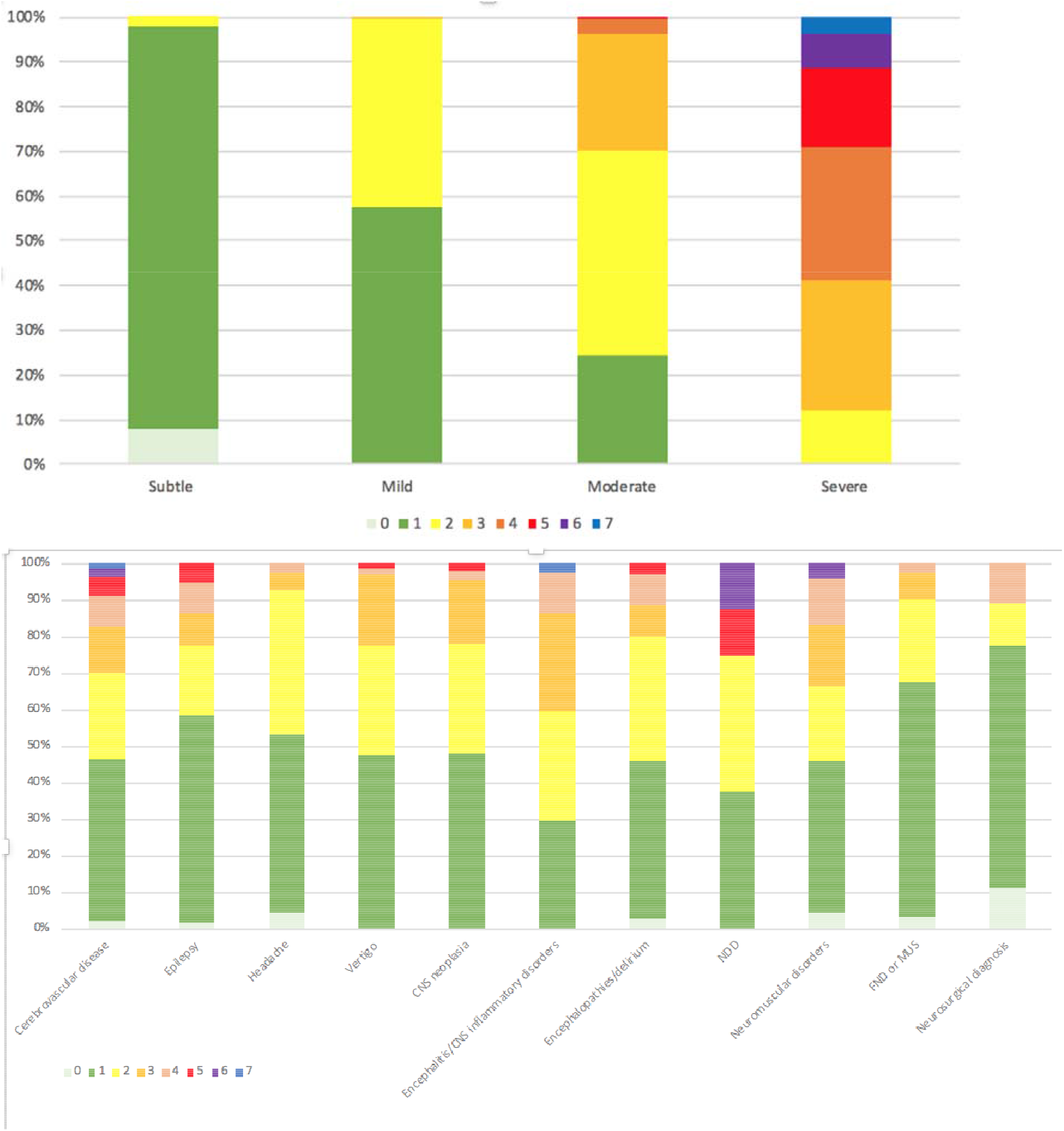
mNIS Complexity Index (mNIS-CI) distribution according to severity classification and within different neurological diagnosis: CNS, central nervous system; FND, functional neurological disorders; ICH, intracerebral hemorrhage; MUS, medically unexplained symptoms; NDD, neurodegenerative disorders.

## DISCUSSION

The study presented a modified version of NIS, including four more items and a composite score and validated its use in a hospital neurology unit for acute inpatients, demonstrating its construct validity, applicability and potential impact in stratifying severity across different neurological conditions.

Standardization of neurological assessment and development of functional rating scales of global impairment are still unmet needs in neurology, both in acute and chronic settings (6,19,20). The NIS was originally developed as a measure of the severity of neurological impairment across a broad range of disabling conditions, including acquired brain injury. The scale was subsequently expanded by adopting a simple ordinal scoring system to capture severity and it has been shown to make a significant contribution to the prediction of functional outcomes in patients with severe/complex disability (11). Although the NIS was considered a useful tool in general neurorehabilitation settings, its psychometric properties, including scalability, reliability, and validity, support the view that it might be a useful measure of neurological impairment both in hospital-based acute and in chronic settings. In this modified version, four new items were included covering important domains which are commonly affected in neurological patients and not covered by the original version, namely *Cranial nerves, Coordination, Involuntary movements*, and *Gait*, giving a total score range of 0–64.

The findings underscore a high correlation between mNIS and NIS scores within each diagnostic group, emphasizing nuanced neurological impairments detected in most patients by both scales. Notably, a substantial proportion of patients (up to one third of the sample) exhibited the newly listed symptoms, highlighting the relevance of these items for a more comprehensive evaluation of neurological severity.

To test construct validity, a significant correlation between total mNIS, NIS, and NIHSS scores were found in patients with Cerebrovascular disease, indicative of consistent severity assessment across scales. Furthermore, specific item-by-item correlations between mNIS, NIHSS, and other cognitive and functional rating scales, showed strong agreements for motor and sensory items and robust correlations for language and visual function assessments.

Of interest, the inter-rater agreement is very high even if the mNIS was carried out by neurology residents, supporting the claim that the mNIS allows reliable scoring by medical staff without a great deal of expertise. Altogether, overall agreement was remarkable, and ratings were highly comparable for most items, even for items such as *Involuntary Movements*, which generally require specific formation, or *such as Mood* and *Other*, greatly relying on individual sensitivity (21,22).

Of note, mNIS total score was significantly associated with functional dependency measures such as BADL, IADL and mRS, supporting the view that mNIS can offer a complementary contribution to the characterization of functional outcomes in a mixed-diagnosis group of patients with acute severe/complex neurological disorders.

Generally, total scores are useful for monitoring clinical course but of little help in stratifying patients according to clinical complexity, due to the different impact of different items and the multiple combinations of cumulative items. We thus developed a Complexity Index which was calculated as the number of categories/domains with sub-items score of 2 or greater. The mNIS-CI correlated with the total score and allowed better identification of complex cases within diagnostic categories and a frequency comparison of complex cases among diagnostic categories.

Some limitations of this study should be considered. First, our sample comprises a large sample of patients admitted to a single-center Neurology unit, thus our findings should be validated a multi-center analysis, across a range of clinical settings and with other samples of neurological patients and physicians (e.g., general practitioners). Second, the scoring process was more reliable for categories in which precise indications had been borrowed from other standardized scales (e.g., NIHSS) and for more objective deficits (e.g., motor impairment) than for patient self-reported deficits (e.g. *Mood*). Thirdly, the validity of mNIS was particularly tested in the Cerebrovascular disease diagnostic category of patients, which was the most numerous and the one evaluated with a widely adopted and known scale such as the NIHSS. Fourth, the study design did not allow us to assess either the clinical utility of the mNIS in terms of long-term outcome, nor the impact of this scale on the clinical decision at discharge.

Nevertheless, our findings applied in a large comprehensive dataset demonstrate that mNIS is a reliable and valid measure of neurological impairment, suitable for being utilized across a broad range of neurological conditions. The mNIS can provide useful information for case-mix adjustment and can assist in the interpretation of clinical outcomes for inpatient with complex neurological diseases.

## CONCLUSION

The mNIS is a valuable tool to measure neurologic severity and complexity in acute inpatients. We confirmed that the mNIS is associated with a high rate of inter-rater agreement, and its domain-specific levels of function are effectively articulated. These results support a wider adoption of the mNIS to assess neurologic function in hospital-based acute settings at different time points, to evaluate its performance in evolving clinical pictures. We suggest that the assessment of neurologic severity by the mNIS might represent a reliable measure either for prognosis or for measuring patients’ outcomes. Furthermore, mNIS complements existing highly valuable disease-specific scales and tools. Finally, mNIS provides standardization for the assessment of neurologic function that will lead to more consistent and accurate description of the neurologic status in terms of severity and complexity. Further efforts will address the translation of the mNIS as a meaningful and reproducible measure of clinical response in different settings.

## Data Availability

http://www.csi.kcl.ac.uk/ukroc

## ACKNOWLEDGEMENTS

The authors gratefully acknowledge the hard work of the Brescia School of Neurology Working Group, and the patients, to whom this work belongs.

## AUTHOR CONTRIBUTIONS

APa: Conceptualization, Formal Analysis, Supervision, Writing Original Draft Preparation – Review & Editing; IM: Data Curation, Formal Analysis, Visualization, Writing – Original Draft Preparation, Writing – Review & Editing; TC: Data Curation, Validation, Visualization; NZ and CZ: Software; MC and EG: Data Curation; AM: Supervision, Writing – Review & Editing; SG and CA: Supervision; LT-S: Writing – Review & Editing; APi: Conceptualization, Data Curation, Formal Analysis, Writing – Original Draft Preparation, Writing – Review & Editing.

## COMPETING INTERESTS

APa received personal compensation as a consultant/scientific advisory board member for Biogen, Lundbeck, Roche, Nutricia, General Healthcare (GE). APa has been supported by grants of the Italian Ministry of University and Research PRIN COCOON (2017MYJ5TH) and PRIN 2021 RePlast (20202THZAW), the H2020 IMI IDEA-FAST (ID853981), Italian Ministry of Health, Grant/Award Number: RF-2018-12366209, RF-2019-12369272 and PNRR-Health PNRR-MAD-2022-12376110. IM, TC, NZ, CZ, MC, EG, SG, AM, CA report no competing interest.

LT-S is Director of the UK Rehabilitation Outcomes Collaborative (UKROC) and was the lead developer of the UKROC tools including the Neurological Impairment Scale. However, neither she nor her employing institution has any financial interest in the tools which are disseminated free of charge.

APi received consultancy/speaker fees from Abbvie, Angelini, Bial, Lundbeck, Roche and Zambon pharmaceuticals and the Movement Disorder Society. APi has been supported by grants of Airalzh Foundation AGYR2021 Life-Bio Grant, The LIMPE-DISMOV Foundation Segala Grant 2021, the Italian Ministry of University and Research PRIN COCOON (2017MYJ5TH), PRIN 2021 RePlast (20202THZAW), PRIN PNRR 2022 ( the H2020 IMI IDEA-FAST (ID853981), Italian Ministry of Health, Grant/Award Number: RF-2018- 12366209 and PNRR-Health PNRR-MAD-2022-12376110.

## FUNDING

No targeted funding reported.

## Notes

### Author Declarations

Ethics committee/IRB of the Brescia Hospital, Brescia, Italy (Neuromultibio study, NP 5285) gave ethical approval for this work

