## Supplemental material for "Time for a standardized neurological assessment in acute setting: the modified Neurological Impairment scale"

Supplementary Figure 1: Flowchart summarizing patients’ selection

mNIS, modified Neurological Impairment Scale; NIHSS, National Institute of Health Stroke Scale

Excluded (n=42): no defined diagnosis at discharge needing clinical follow-up

Excluded (n=122): missing data for multidimensional comorbidity/frailty/disability evaluation

1081 patients with definite diagnosis and complete multidimensional evaluation at admission and discharge

1245 patients admitted from January-December 2023

40 patients with mNIS evaluation performed by two different raters (inter-rater agreement)

1081 patients with complete multidimensional assessment at admission and discharge

Supplementary Table 1: Clinical Characteristics of the cohort

| Evaluation at admission | Adult (<65 y) | Elderly (≥65 y) |
| --- | --- | --- |
|  | 454 | 627 |
| Age, mean ± SD | 47.26±14.17 | 77.74±7.00 |
| Sex, female n% | 47.8 | 51.4 |
| Comorbidities |  |  |
| CIRS cumulative severity, mean±SD | 1.37 ± 1.52 | 3.18 ± 2.10 |
| CIRS number of comorbidities 0-1, n% | 62.8 | 21.6 |
| 2-3, n% | 27.1 | 40.9 |
| 4-6, n% | 9.2 | 31.0 |
| > 6, n% | 0.8 | 6.8 |
| mRS premorbid, mean (± SD | 0.78 ± 1.07 | 1.52 ± 1.40 |
| Normal (mRS 0), n% | 51.8 | 27.0 |
| Mild disability (mRS 1-2), n% | 40.7 | 48.9 |
| Moderate/severe disability (mRS 3-5), n% | 7.5 | 24.1 |

CIRS, Cumulative Illness Rating scale; mRS, modified Rankin scale; SD, standard deviation; y, years old.

Supplementary Table 2: Cross-validity of mNIS items

| **Item mNIS** | **Item cross-valid** | **r** | **p*** | **n** |
| --- | --- | --- | --- | --- |
| Left arm | Left arm, NIHSS | 0.79 | 0.001 | 435 |
| Right arm | Right arm, NIHSS | 0.81 | 0.001 | 435 |
| Left leg | Left leg, NIHSS | 0.81 | 0.001 | 435 |
| Right leg | Right leg, NIHSS | 0.81 | 0.001 | 435 |
| Sensitive deficits | Sensitive deficits, NIHSS | 0.70 | 0.001 | 435 |
| Language | Aphasia, NIHSS | 0.76 | 0.001 | 435 |
| Language | Dysarthria, NIHSS | 0.47 | 0.001 | 435 |
| Visual functions | Gaze, NIHSS | 0.43 | 0.001 | 435 |
|  | Visual field, NIHSS | 0.67 | 0.001 | 435 |
| Neglect | Neglect, NIHSS | 0.64 | 0.001 | 435 |
| Mental status | Consciousness, NIHSS | 0.48 | 0.001 | 435 |
| Mental status | Orientation, NIHSS | 0.59 | 0.001 | 435 |
| Mental status | Comprehension, NIHSS | 0.47 | 0.001 | 435 |
| Mental status | MMSE, total score | 0.61 | 0.001 | 62 |
| Mental status | CAM, total score | 0.46 | 0.001 | 433 |
| Cranial nerves | VII cranial nerve deficit, NIHSS | 0.70 | 0.001 | 435 |
| Coordination | Ataxia, NIHSS | 0.77 | 0.001 | 435 |
| Gait | Tinetti, total score | 0.97 | 0.001 | 435 |

* Correlations between mNIS items and items able to quantify the same function/domain from other functional scales

NIHSS, NIH stroke scale; MMSE, MiniMental State Examination; CAM, confusion assessment method.

Supplementary Table 3: Inter-rater agreement for mNIS (total and sub scores).

| **Item** | **Cronbach’s alpha** | **ICC (95% CI)** |
| --- | --- | --- |
| Motor deficits | 0.87 | 0.76 (0.60-0.87) |
| Sensitive deficits | 0.75 | 0.60 (0.36-0.77) |
| Language | 0.90 | 0.82 (0.69-0.90) |
| Tone | 0.65 | 0.60 (0.36-0.77) |
| Visual functions | 0.66 | 0.49 (0.21-0.69) |
| Neglect | 0.64 | 0.45 (0.18-0.68) |
| Mental status | 0.79 | 0.66 (0.44-0.80) |
| Hearing | 0.79 | 0.66 (0.44-0.80) |
| Pain | 0.79 | 0.65 (0.43-0.80) |
| Behavioural alterations | 0.77 | 0.63 (0.40-0.79) |
| Mood | 0.49 | 0.33 (0.02-0.58) |
| Fatigue | 0.73 | 0.57 (0.32-0.75) |
| Other | 0.48 | 0.32 (0.01-0.57) |
| **Added items** |  |  |
| Cranial nerves | 0.87 | 0.77 (0.61-0.87) |
| Coordination | 0.71 | 0.55 (0.29-0.73) |
| Involuntary movements | 0.51 | 0.34 (0.04-0.59) |
| Gait | 0.81 | 0.68 (0.48-0.82) |
| **Total NIS** | **0.93** | **0.87 (0.77–0.93)** |
| **Total mNIS** | **0.95** | **0.90 (0.82-0.95)** |

IIC, intraclass correlation coefficient; (m)NIS, (modified) neurological impairment scale.
